# A proteome-wide genetic investigation identifies several SARS-CoV-2-exploited host targets of clinical relevance

**DOI:** 10.1101/2021.03.15.21253625

**Authors:** Mohd A Karim, Jarrod Shilts, Jeremy Schwartzentruber, James Hayhurst, Annalisa Buniello, Elmutaz Shaikho Elhaj Mohammed, Jie Zheng, Michael V Holmes, David Ochoa, Miguel Carmona, Joseph Maranville, Tom R. Gaunt, Valur Emilsson, Vilmundur Gudnason, Ellen M. McDonagh, Gavin J. Wright, Maya Ghoussaini, Ian Dunham

## Abstract

The virus SARS-CoV-2 can exploit biological vulnerabilities in susceptible hosts that predispose to development of severe COVID-19. Previous reports have identified several host proteins related to the interferon response (e.g. OAS1), interleukin-6 signalling (IL-6R), and the coagulation cascade (linked via ABO) that were associated with risk of COVID-19. In the present study, we performed proteome-wide genetic colocalisation tests leveraging publicly available protein and COVID-19 datasets, to identify additional proteins that may contribute to COVID-19 risk. Our analytic approach identified several known targets (e.g. ABO, OAS1), but also nominated new proteins such as soluble FAS (colocalisation probability > 0.9, p = 1 × 10^−4^), implicating FAS-mediated apoptosis as a potential target for COVID-19 risk. We also undertook polygenic (pan) and cis-Mendelian randomisation analyses that showed consistent associations of genetically predicted ABO protein with several COVID-19 phenotypes. The ABO signal was associated with plasma concentrations of several proteins, with the strongest association observed with CD209 in several proteomic datasets. We demonstrated experimentally that CD209 directly interacts with the spike protein of SARS-CoV-2, suggesting a mechanism that could explain the ABO association with COVID-19. Our work provides a prioritised list of host targets potentially exploited by SARS-CoV-2 and is a precursor for further research on CD209 and FAS as therapeutically tractable targets for COVID-19.

## Introduction

At the current time, the coronavirus disease 2019 (COVID-19) pandemic is implicated in deaths of more than two million people worldwide^1^. Fortunately, effective vaccines have been deployed and are expected to substantially reduce mortality and morbidity due to severe COVID-19. However, several mutated strains of SARS-CoV-2 virus have emerged that are reported to be more contagious than the original strain^2,3^ and it is currently unclear to what extent the vaccines will be efficacious against the mutated strains^4^. It is likely that the mutated strains of SARS-CoV-2 will continue to exploit the same vulnerable host biology and, in susceptible individuals, evade immune defenses and promote the excessive host inflammatory response that is characteristic of severe COVID-19 disease^5^. Therefore, identification of host proteins that play roles in COVID-19 susceptibility and severity remain crucial to the development of therapeutics as host protein mechanisms are independent of genomic mutations in the virus. In some cases, such therapeutics may also be important in tackling viruses beyond SARS-CoV-2^6^.

Several large-scale systematic experimental efforts have identified key host proteins that interact with viral proteins in the pathogenesis of severe COVID-19^5,7,8^. To complement *in vitro* host protein characterisation efforts, several groups have leveraged genetic datasets of human proteins and COVID-19 disease to identify therapeutically actionable candidate host proteins that are likely to play roles in enhancing COVID-19 susceptibility or to be involved in the pathogenesis of severe COVID-19^9,10^. One of the approaches used was Mendelian randomisation (MR). MR simulates the design of randomised trials, with the underlying principle that randomisation of alleles at conception offers the opportunity to examine approximate differences in average risk of a disease between comparable groups in a population that differ only in the distribution of the risk factor of interest^11^, for example protein abundance^12^. This allows use of alleles as genetic instruments representing genetically predicted protein levels to proxy effects of pharmacological modulation of the protein. Some of the clinically actionable proteins identified by the MR approach are part of type I interferon signalling (encoded by genes: *IFNAR2, TYK2, OAS1*) and interleukin-6 (IL-6) signalling pathways (*IL6R*). Only one of these proteins (encoded by *OAS1*) had any evidence of genetic colocalisation, i.e. evidence that genetic associations of the protein and COVID outcomes shared the same causal genetic signal^10^. An additional protein that was supported by both MR and genetic colocalisation tests was ABO^10^, reported in several published genome-wide association studies (GWAS) of COVID-19^9,13^. In response to the first published GWAS of COVID-19, we reported findings that link the *ABO* signal with a number of clinical actionable targets including coagulation factors (von Willebrand factor, vWF and Factor VIII, F8), IL-6, and CD209/DC-SIGN^14^.

However, in most of the previous MR studies^9,10^, investigators only used curated cis-acting variants (genetic variants near or in the gene encoding the relevant protein) as genetic instruments to represent effects of genetically predicted protein concentrations, rather than genome-wide instruments. While use of cis-acting variants can minimise the risk of horizontal pleiotropic effects (i.e. associations driven by other proteins not on the causal pathway for the disease), it can suffer from lower power than a genome-wide analysis due to fewer available instruments^12^. Furthermore, in previous protein-COVID-19 MR studies, genetic colocalisation tests were carried out only for protein-phenotype associations that were significant in the MR analysis, potentially excluding many protein-phenotype associations that may share the same causal genetic signal but are underpowered in a proteome-wide MR approach.

In the present study, we expanded on these previous reports by undertaking a proteome-wide two-sample pan- and cis-MR analysis using the Sun et al. GWAS^15^ of plasma protein concentrations and several COVID-19 GWAS phenotypes from the ICDA COVID-19 Host Genetics Initiative (Oct. 2020 release)^16^. First, we showed that genetically-predicted circulating ABO protein was associated with COVID-19 susceptibility and severity and the lead *ABO* signal was associated strongly with plasma concentrations of soluble CD209. Second, we collected evidence for a direct mechanism of interaction between the SARS-CoV-2 spike protein and human CD209 protein. Third, we performed proteome-wide genetic colocalisation tests, followed by single-instrument cis-MR analysis, and we report additional novel targets of therapeutic relevance. Finally, we examined associated phenotypes using the colocalising signals from the Open Targets Genetics portal (http://genetics.opentargets.org) to shed light on the biological basis of association of the proteins with the COVID-19 phenotypes.

## Methods

### Genetic associations of proteins

We primarily used Sun et al protein GWAS data^15,17^ for the pan-/cis-MR analyses and for performing genetic colocalisation tests (described below). The pan-/cis-MR effects were expressed per standard deviation (SD) higher genetically predicted plasma protein concentrations. Two additional proteomic datasets^17,18^ were used to identify proteins associated with the *ABO* locus. The genotyping protocols and QC of these proteomic studies have been described previously^15,17,18^. All three of the proteomic studies have used the SOMAscan assay platform (an aptamer-based protein detection platform) to detect and quantify protein abundance^19^.

### Genetic associations of COVID-19

We used seven meta-analysed COVID-19 data sets from the October 2020 release of the ICDA COVID-HGI group (https://www.covid19hg.org/results/r4/). These seven COVID-19 outcomes are A1 (Very severe respiratory confirmed covid vs. not hospitalized covid), A2 (Very severe respiratory confirmed covid vs. population), B1 (Hospitalized covid vs. not hospitalized covid), B2 (Hospitalized covid vs. population), C1 (Covid vs. lab/self-reported negative), C2 (Covid vs. population), and D1 (Predicted covid from self-reported symptoms vs. predicted or self-reported non-covid). Definitions of these outcomes are provided in the supplement table **Table S1**.

### Harmonisation of protein and COVID summary statistics

Prior to analyses, we performed a liftover of datasets that reported genomic coordinates using the GRCh37 assembly to GRCh38. We also checked and ensured that the effect allele in a GWAS locus is the alternative allele in the forward strand of the reference genome. To infer strand for palindromic variants (variants with A/T or G/C alleles, i.e. variants with the same pair of letters on the forward strand as on the reverse strand), we first checked the orientation of all non-palindromic variants with respect to the reference genome to assess whether there was a strand consensus of 99% or more. For example, for a given GWAS, if ≥ 99% of the non-palindromic variants were on the forward strand, we assumed the palindromic variant would also be on the forward strand, otherwise they were excluded from analyses. Details of the harmonisation workflow are provided in our github pages^20,21^.

### Mendelian randomization

To construct genetic instruments for MR analysis, we selected near-independent (r^2^ = 0.05) genetic variants from across the genome (‘*pan*’-instruments) or from within +1 Mbp from transcription start site (TSS) of the gene encoding the protein (‘*cis*’-instruments) associated with the encoded protein abundance at p <= 5 × 10^−8^ for pan-MR analyses and at a less stringent p <= 1 × 10^−5^ for cis-MR analyses (this p-value corrects for the number of proteins in the druggable genome^22^). We used the generalised summary data-based Mendelian randomization (GSMR) approach with the heterogeneity independent instrument (HEIDI)-outlier flag turned on to carry out the pan- and cis-MR analyses^23^. The GSMR software, using the HEIDI-outlier method, removes potentially pleiotropic instruments and accounts for residual correlation between instruments (important as we are using near-independent genetic instruments). For each COVID-19 outcome, we used the Benjamini-Hochberg FDR threshold of 5% for significance. Genotype data from 10,000 randomly sampled UK Biobank participants was used as a reference panel. For trans-acting instruments in pan-MR associations, variants were mapped to their respective cis-gene that had the highest overall V2G score in the Open Targets Genetics portal^24–26^.

### Colocalisation analysis and phenome-wide association study

To identify shared causal genetic signals between protein and COVID outcomes, we used the Bayesian method of genetic colocalisation implemented in the *coloc* R package^27^, assuming one independent signal in each region. We used beta and standard errors of cis-pQTLs of phenotype pairs as inputs. The default priors in *coloc* were used, i.e. the prior of either genetic signal is 1 × 10^−4^, and the prior of two genetic signals being shared is 1 × 10^−5^. For each COVID-19 outcome, a posterior probability for shared causal genetic signal (PP.H4) threshold more than 0.8 was used to identify shared causal genetic variants. Using the colocalising signals, we carried out a phenome-wide association study (PheWAS) using phenotype data (n = ∼ 3000 GWAS) from the Open Targets Genetics portal^24,25^.

### Evidence against aptamer binding artefacts

To identify variants associated with proteins due to aptamer or epitope binding artefacts^28^, we assessed whether genetic instruments for MR or coloc-based single-SNP MR analysis were associated with corresponding gene expression (i.e. whether they were also cis-eQTLs). This used gene expression data from the Open Targets Genetics portal^24^. SNPs that were not cis-eQTLs were investigated further by identifying whether they were (or were in linkage disequilibrium or LD at *r*^*2*^ = 0.8 with) missense variants. To query if variants were missense or in LD with missense variants, we used the *LDlinkR* R package with LD data from the 1000 genome phase 3 EUR population^29^. Where cis-pQTLs were not cis-eQTLs and were missense variants (or in LD with missense variants at *r*^*2*^ = 0.8) affecting the respective genes, these proteins were flagged and excluded from any further downstream analyses on the basis that the missense variant(s) might influence aptamer binding. Where cis-pQTLs were also cis-eQTLs and were missense variants (or in LD with missense variants) for the respective genes, although the effect estimates would not be valid, the causal inference using the instruments are unlikely to be biased; hence, these variants were retained in supplementary tables and estimates of probes represented by these variants were flagged in the main figures. In the rest, where cis-pQTLs had an effect on gene expression but were not missense variants or in LD with missense variants, were included in all analyses and presented without restrictions.

### Recombinant protein production

Recombinant human receptors and SARS-CoV-2 spike protein extracellular domains were expressed and purified as previously described^30^. Briefly, the full extracellular domain sequences of each were expressed as soluble secreted proteins in HEK293 cells. All proteins were affinity-purified using their hexahistidine tags. For biotinylated proteins, co-transfection of secreted BirA ligase in the presence of 100 µM D-biotin resulted in the covalent addition of a biotin group to an acceptor peptide tag, also as described previously^31^. The extracellular domain of CD209 (Q9NNX6) was defined as beginning at Pro114, while the full cDNA sequence was acquired from OriGene (#SC304915).

### Plate-based protein binding assay

The binding of biotinylated human receptor extracellular domains to pentameric SARS-CoV-2 spike protein was measured using the avidity-based extracellular interaction assay (AVEXIS) as previously described^32^. Briefly, the wells of a streptavidin-coated 96-well plate were saturated with biotinylated bait of either CD209, ACE2, or a previously-described negative-control construct consisting only of the C-terminal protein tags shared by all other recombinant proteins (rat Cd4(d3+4)-linker-Bio-6xHis)^33,34^. Across these baits we applied a dilution series of the full SARS-CoV-2 spike protein extracellular domain pentamerized by a peptide sequence from the cartilage oligomeric matrix protein with a beta lactamase reporter. After washing, binding was measured by hydrolysis of a colourimetric nitrocefin substrate whose product was quantified by light absorbance at 450nm.

### Cell-based receptor binding assay

HEK293 cells were transiently transfected as described previously^35^ with expression plasmids encoding full-length cDNA of CD209 (Origene SC304915), or a mock transfection lacking the expression plasmid. Separately, recombinant biotinylated spike protein was tetramerized around streptavidin conjugated to phycoerythrin as previously described^36^. Cells were incubated with tetramers of spike or a control construct of protein tags before being analyzed on a flow-cytometer as previously described^30^.

## Data availability

Summary data used for genetic analyses are publicly available (Sun et al can be downloaded from GWAS catalog https://www.ebi.ac.uk/gwas/downloads/summary-statistics and COVID-19 HGI summary statistics can be downloaded from their website https://www.covid19hg.org/results/). Data generated from our study are provided in the supplementary tables (pan-MR and cis-MR association results filtered at p < 0.05 and no filters applied to colocalisation results).

## Code availability

Codes used to harmonise summary statistics are provided in https://github.com/EBISPOT/gwas-sumstats-harmoniser. Codes for pan- and cis-MR analyses are provided in the GSMR website (https://cnsgenomics.com/software/gcta/#GSMR). Codes for genetic colocalisation analyses are provided in the coloc github page (https://github.com/chr1swallace/coloc). All codes used in the paper to reproduce results are provided in https://github.com/mohdkarim/covid_paper.

## Results

### Pan- and cis-MR analyses support a role of circulating ABO protein concentrations and soluble IL-6R in COVID-19 risk

Our multi-instrument MR analysis used both genetic variants from across the genome (pan-MR) and genetic variants near or in the gene encoding the relevant protein (cis-MR) to investigate associations of genetically predicted plasma protein concentrations with risk of COVID-19 outcomes. The COVID-19 outcome definitions are provided in **Table S1**. Although the pan-MR analysis leveraged genetic data from both cis- and trans-acting pQTLs (with selection of pQTLs from across the genome automated by GSMR’s built-in HEIDI-outlier exclusion method), for some protein-COVID-19 pairs that were associated at 5% FDR, the associations with COVID-19 outcomes were exclusively driven by trans-acting pQTLs or cis-acting genetic instruments. For example, although 6 proteins were represented by both cis- and trans-acting genetic instruments, 2 (ABO and IL6R) were represented only by cis-acting variants, and 1 (SELE) was driven entirely by trans-acting instruments (mainly *ABO* trans-pQTLs) (**Table S2**). Overall, the pan-MR analysis revealed 9 distinct protein probes associated with 4 COVID outcomes at an FDR of 5% (**Figure 1A**). The pQTLs selected by GSMR to represent these 9 probes were also cis-eQTLs (as curated for the Open Targets Genetics portal^37^) and were not missense variants or in LD with missense variants (**Table S3**), minimising the possibility that SNPs with artefactual associations with proteins were used as genetic instruments.

**Figure 1:**
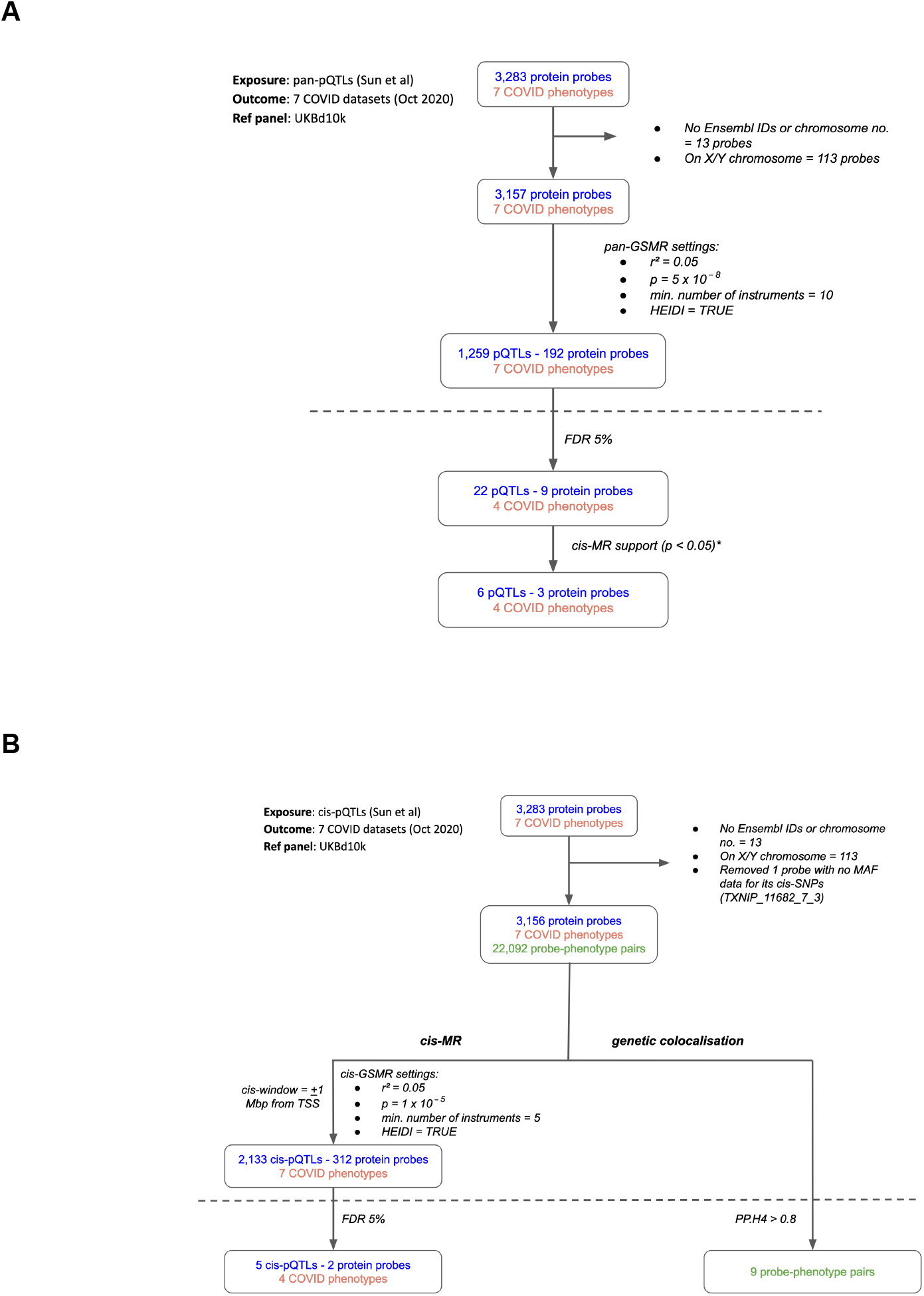
Flowcharts illustrating the process of A. pan-MR, and B. cis-MR and genetic colocalisation. Both pan and cis-MR methods used Sun et al.^15^ as the source of genetic instruments and the UK Biobank downsampled 10k (UKBd10k) individual genotype data as reference panel. We selected near-independent genetic instruments and performed two sample MR analysis using GSMR that adjusted for residual correlation between instruments. Genetic colocalisation analysis was used to estimate posterior probabilities of shared causal genetic signal between protein and outcomes. A posterior probability of shared causal genetic signal of more than 0.6 (i.e. a PP.H4 or posterior probability for hypothesis four > 0.6) was used as evidence of genetic colocalisation. The dashed line separates analysis (above the line) from target curation (below the line). *Only three proteins with pan-MR evidence of association with COVID also had cis-MR evidence support at nominal cis-MR p-value < 0.05.

While the pan-MR analysis used genetic data from across the genome, the cis-MR analysis restricted genetic instrument selection to those near (within 1 Mb of TSS) or in the gene encoding the protein. Three proteins with pan-MR associations were supported by corresponding cis-MR associations (**Figure 1A, 1B, Table S4**): ABO, ICAM-1, and IL-6R. Among these three, only ABO and IL-6R proteins had some evidence of genetic colocalisation with posterior probabilities (PP.H4) more than 0.9 and 0.4, respectively, of a shared genetic signal between protein and COVID-19 phenotype (**Figure 2**). Although the PP.H4 of IL-6R was very weak (0.4), it had a positive (H4/H3 = 3.6) indicating a common signal of the IL-6R protein with the COVID-19 outcome is a more likely scenario than the association driven by two independent signals.

**Figure 2:**
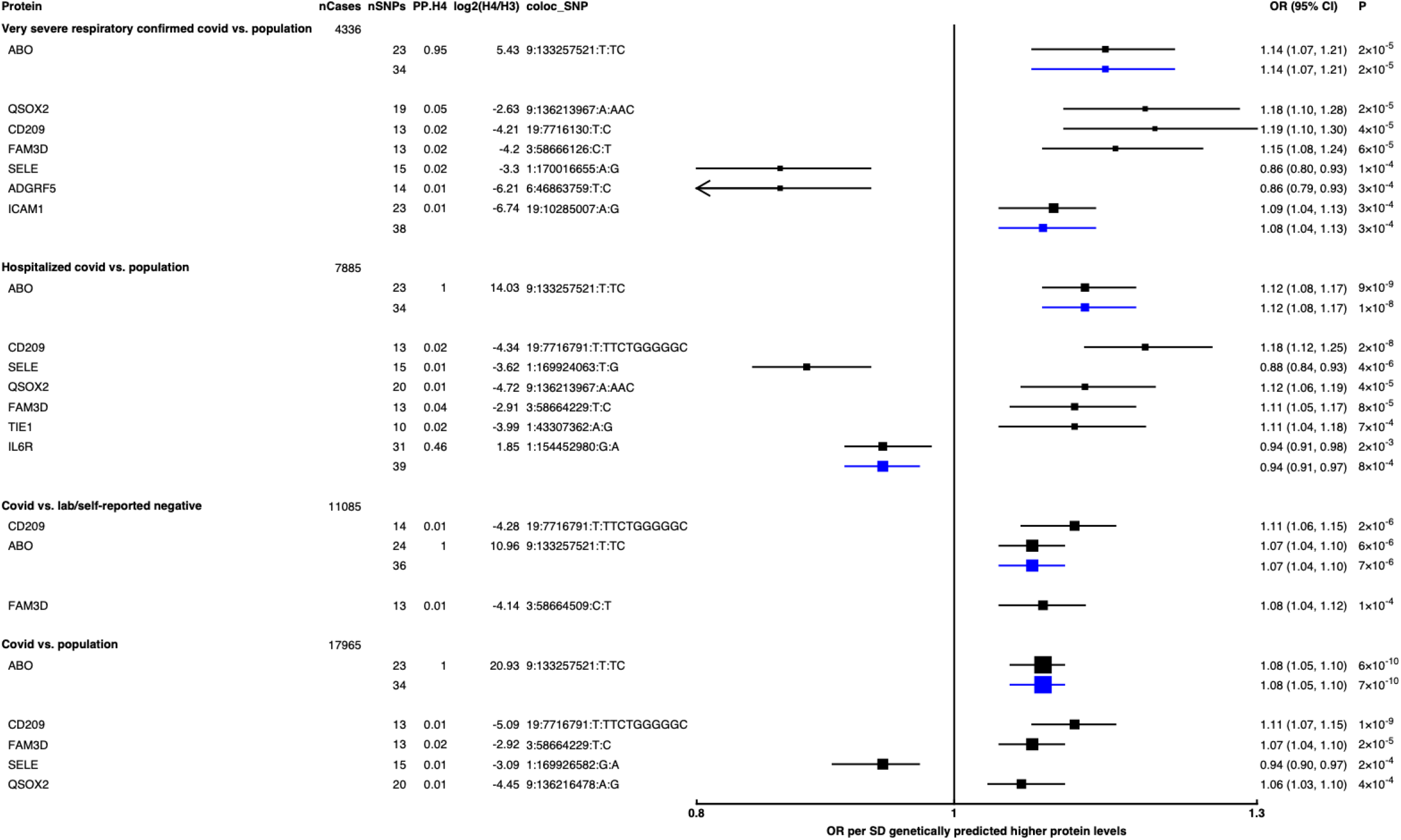
Forest plot illustrating associations of genetically predicted plasma protein concentrations with selected COVID-19 phenotypes. The black point estimates represent odds ratios (OR) of COVID-19 outcome per SD increase of genetically predicted protein abundance using genetic instruments from across the genome (pan-MR). The blue point estimates represent OR of COVID outcome per SD increase of genetically predicted protein abundance using genetic instruments near or in the gene encoding the protein (cis-MR). Error bars represent 95% confidence intervals (95% CI). The areas of the squares are proportional to the inverse of the variance of the log ORs. For each COVID phenotype, pan-MR associations at FDR 5% were retained. Each row under a COVID phenotype represents a pQTL and includes the number of cases in the COVID phenotype (nCases), the number of SNPs used as genetic instruments for the protein (nSNPs), the posterior probability that protein and COVID traits colocalise (PP.H4), the posterior probability evidence for vs. against shared causal variants (log2(H4/H3)), and the candidate colocalising signal (coloc_SNP).

Genetically predicted ABO concentration was associated with risk in 4 out of 7 COVID-19 outcomes (**Figure 2**). These 4 outcomes represented both susceptibility (e.g. COVID-19 vs. population, cis-MR odds ratio (OR) [95% CI] per SD genetically predicted ABO concentrations: 1.08 [1.05, 1.10], p = 7 × 10^−10^) and severity (e.g. hospitalised COVID-19 vs. population, cis-MR OR [95% CI]: 1.12 [1.08, 1.17], p = 1 × 10^−8^) of COVID-19. Genetically predicted soluble IL-6R was only associated with higher risk of hospitalised COVID-19 compared to population-based controls (cis-MR OR [95% CI] per SD genetically predicted IL-6R: 0.94 [0.91, 0.97], p = 8 × 10^−4^) (**Figure 2)**.

When examining the SNPs involved in the pan-MR associations of the 9 probes, all probes except IL-6R and ABO had at least one trans-acting SNP and in all these cases, at least one of the trans-acting SNPs were assigned to the *ABO* gene by the Open Targets Genetics V2G pipeline (**Table 1**), re-confirming the pervasive pleiotropy of the *ABO* genetic signal. Furthermore, when examining the consistency of pan-MR associations of these 9 probes across all 7 COVID-19 outcomes, the protein probes that have trans-acting *ABO* SNPs exhibited a similar association profile as the ABO protein probe, associated with only COVID-19 outcomes that have population-based controls (**Table S5**).

**Table 1:** Summary of proteins reported in our study and the different sources of evidence supporting their prioritization. *Where trans-acting SNPs are used, genes assigned to SNPs with the highest Variant-to-Gene scores in Open Targets Genetics were used for annotation. GC: genetic colocalisation; MR: Mendelian randomisation.

| Protein | Supported by multi-instrument pan-MR |  |  | Supported by multi-instrument cis-MR | Supported by GC & single-SNP cis-MR | Experimental support | Existing drugs | Previously reported |
| --- | --- | --- | --- | --- | --- | --- | --- | --- |
|  | No. of cis-acting SNPs | No. of trans-acting SNPs | trans-acting gene(s)* |  |  |  |  |  |
| **ABO** | 93 | 0 | None | ✓ | ✓ | x | x | ✓ |
| **QSOX2** | 16 | 63 | *ABO, OBP2B, ADAMTS13* | x | x | x | x | x |
| **CD209** | 8 | 45 | *ABO, SURF6* | x | x | ✓ | x | ✓ |
| **FAM3D** | 8 | 44 | *ABO, SULT2B1, FAM83E, NTN5, FUT2* | x | x | x | x | x |
| **SELE** | 0 | 60 | *ABO, FAM118B, RALGDS, OBP2B, ADAMTS13, SURF1* | x | x | x | ✓ | x |
| **ADGRF5** | 5 | 9 | *ABO, IL6ST, ADAMTS13* | x | x | x | x | x |
| **ICAM1** | 71 | 1 | *ABO* | ✓ | x | x | ✓ | x |
| **TIE1** | 2 | 18 | *ABO, ST3GAL6, GBGT1, SURF6* | x | x | x | x | x |
| **IL6R** | 62 | 0 | None | ✓ | x | x | ✓ | ✓ |
| **FAS** | No | No | No | x | ✓ | x | x | x |
| **OAS1** | No | No | No | x | ✓ | x | x | ✓ |
| **THBS3** | No | No | No | x | ✓ | x | x | x |

### CD209/DC-SIGN: a proposed alternate entry receptor for SARS-CoV-2

The *ABO* signal (rs8176719-TC) overlaps several genes. We explored proteome-wide associations in three separate proteomic datasets^15,18,38^. Aside from the ABO protein, the *ABO* SNP rs8176719-TC showed the strongest association (Sun et al: p = 6.03 × 10^−258^, Emilsson et al: p = 1.00 × 10^−307^, Suhre et al: p = 1.27 × 10^−75^) with higher plasma concentrations of soluble CD209 in all three datasets (associations from two datasets illustrated in **Figure 3**, and associations from all three datasets tabulated in **Table S6**). To validate this as a relevant target for COVID-19, we experimentally tested whether CD209 directly interacts with SARS-CoV-2, as had been recently proposed. We found that the full-length ectodomain of the SARS-CoV-2 spike protein specifically bound to human CD209, both in purified recombinant forms and on the surface of human cells expressing CD209 receptors (**Figure 4A, 4B**).

**Figure 3:**
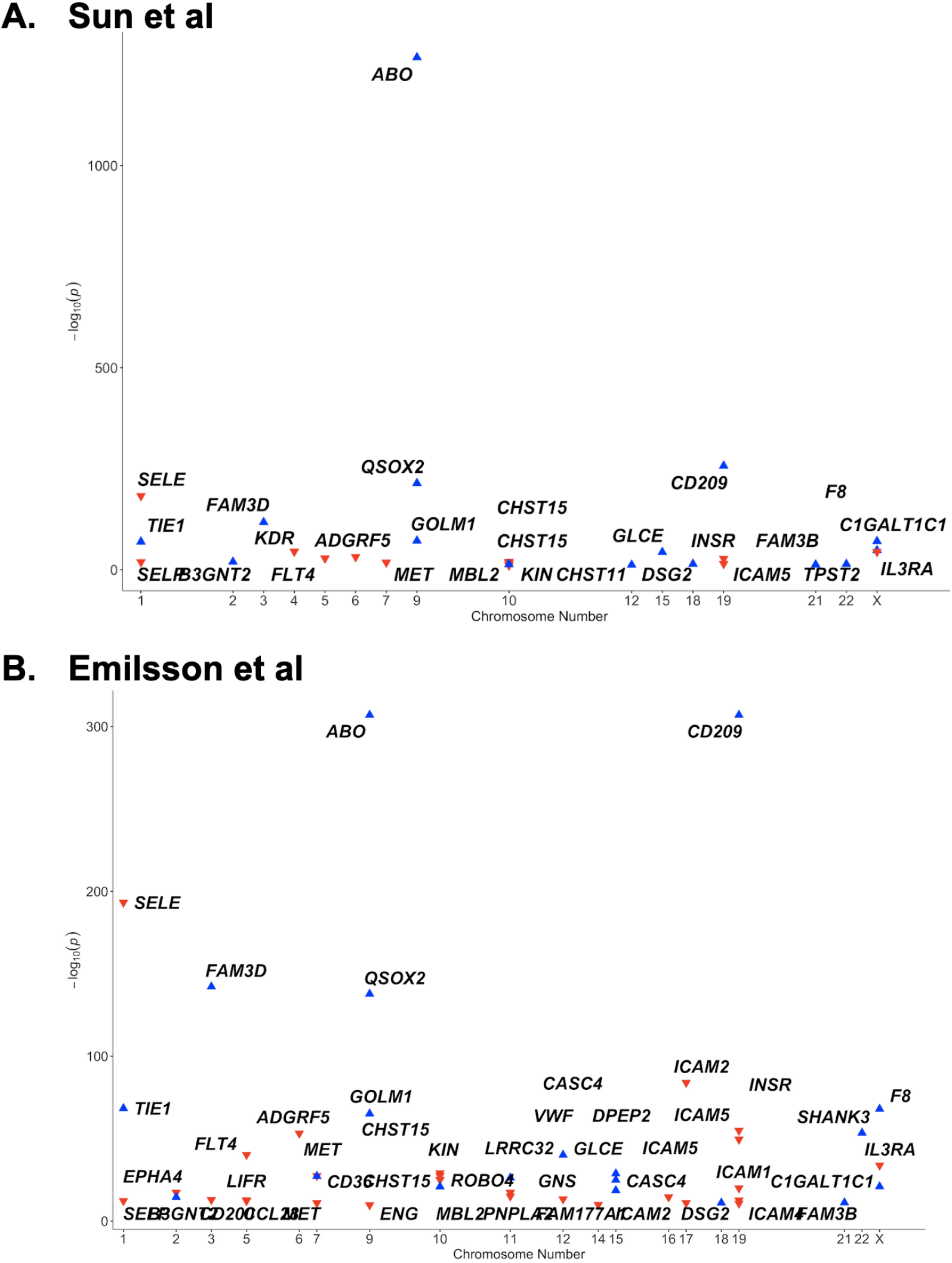
Proteome-wide association of the *ABO* signal (rs8176719-TC) in A. Sun et al, and B. Emilsson et al datasets. The x-axis represents the chromosome for the gene encoding the protein. The y-axis represents the p-value of the per-allele association of rs8176719-TC (or a SNP in high linkage disequilibrium at r2 > 0.8 with rs8176719-TC) with the proteins in Sun et al and Emilsson et al datasets. The red triangles point downwards and denote the inverse association of the *ABO* signal with the protein. The blue triangles point upwards and denote the positive association of the *ABO* signal with the protein. Only proteins that were considered significant at the study-specific Bonferroni-corrected p-value thresholds are displayed in this plot and tabulated in **Table S6** (**Table S6** also reports associations from an additional protein dataset - Suhre et al^18^).

**Figure 4:**
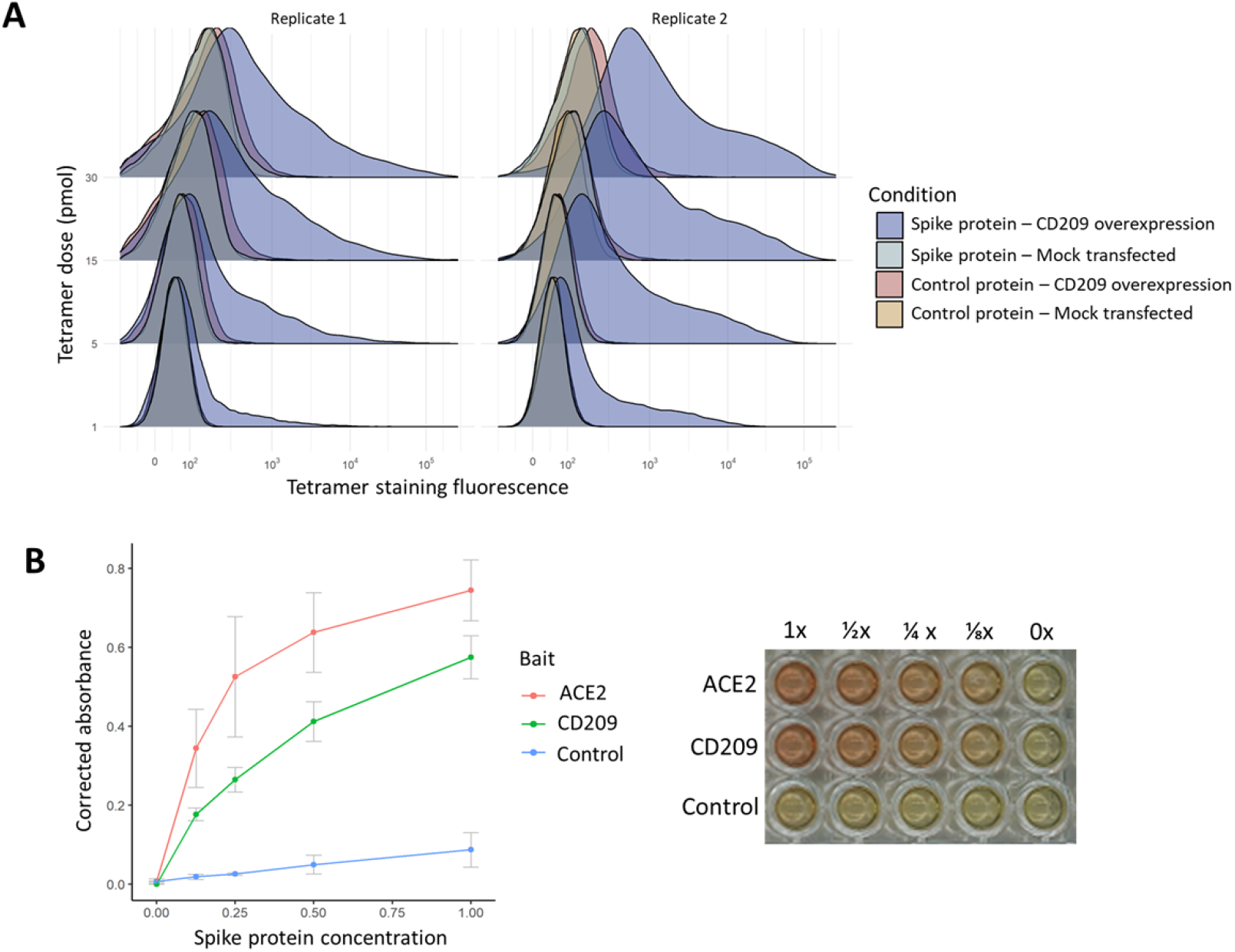
*In vitro* binding experiments with purified SARS-CoV-2 spike protein confirm human CD209 as a functional binding target. A. Human cell lines overexpressing cell-surface CD209 protein gain the ability to specifically bind SARS-CoV-2 spike. The density plots represent flow cytometry measurements of HEK293 cells stained with fluorescently-conjugated tetramers of SARS-CoV-2 spike protein or a tag-only protein control. Blue distributions are cells with surface CD209, while red are control-transfected cells. Light shades indicate a negative control tetramer that was used for staining, while dark shades are stained with spike protein. **B**. Purified recombinant CD209 ectodomains interact with the spike protein of SARS-CoV-2 in an *in vitro* binding assay. A dilution series of purified spike protein was applied over immobilized CD209, ACE2 (positive control) or a negative control protein. A plot of quantified absorbance is displayed alongside a representative assay plate. Error bars are standard deviations of 2 replicates.

### Proteome-wide genetic colocalisation implicates additional proteins in COVID-19 risk including FAS, SCARA5, and OAS1

To identify additional proteins associated with risk of COVID-19, we conducted proteome-wide genetic colocalisation tests followed by single-SNP MR analysis (**Table S7**). This ‘coloc-first’ approach identified 4 proteins (ABO, FAS, OAS1, THBS3) with evidence of genetic colocalisation (PP.H4 > 0.8) with 4 out of 7 COVID-19 phenotypes (**Figure 5**). Two of these (FAS and THBS3) are, to the best of our knowledge, not reported in proteomic MR studies of COVID-19 to date which have only examined for colocalisation evidence after MR.

**Figure 5:**
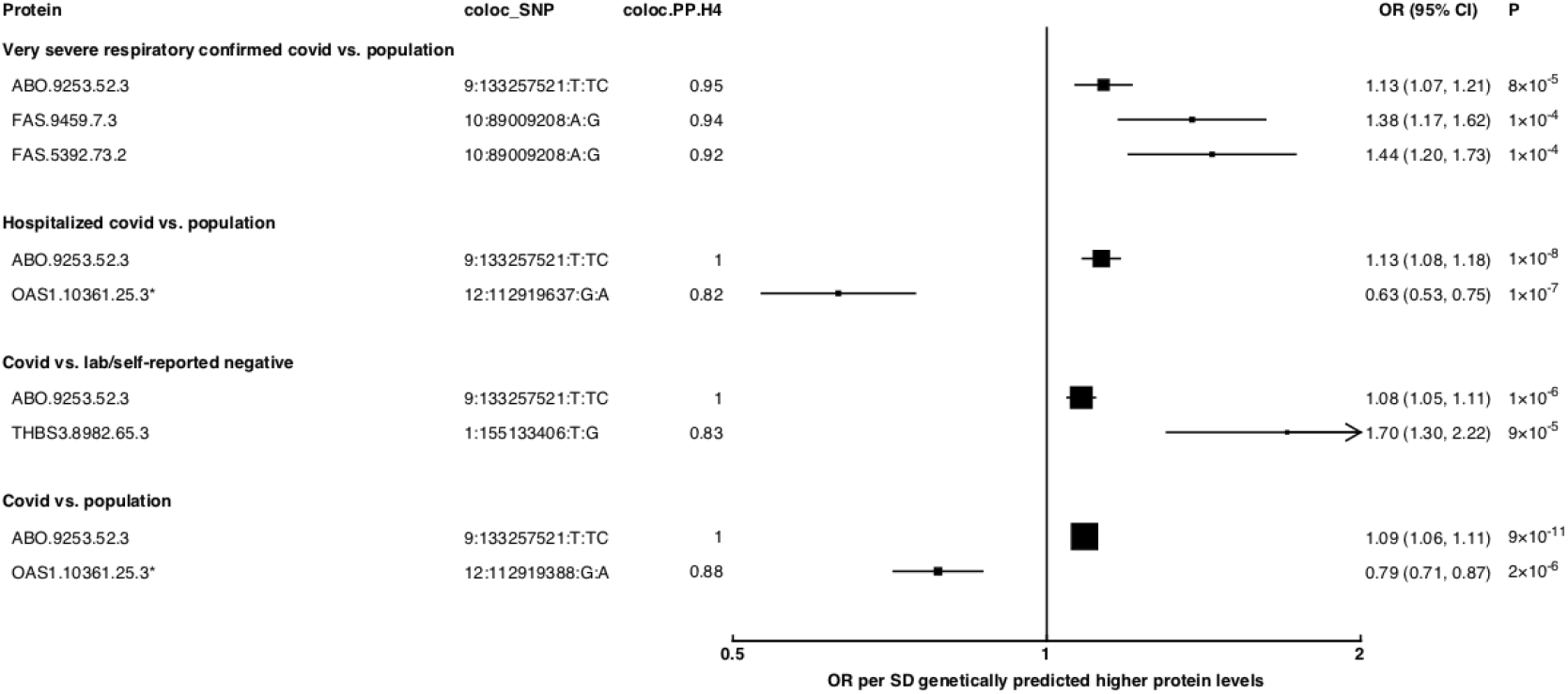
Forest plot illustrating associations of genetically predicted plasma protein concentrations that colocalised with the selected COVID-19 phenotypes (PP.H4 > 0.6). The black point estimates represent odds ratios (OR) of COVID-19 outcome per SD increase of genetically predicted protein abundance using single-SNP colocalising signals (coloc_SNP). Error bars represent the 95% confidence interval around the estimates. The areas of the squares are proportional to the inverse of the variance of the log ORs. *Denotes proteins that have coloc_SNP that are either missense variants or in LD with missense variants, rendering their effect estimates potentially biased.

**Figure 6:**
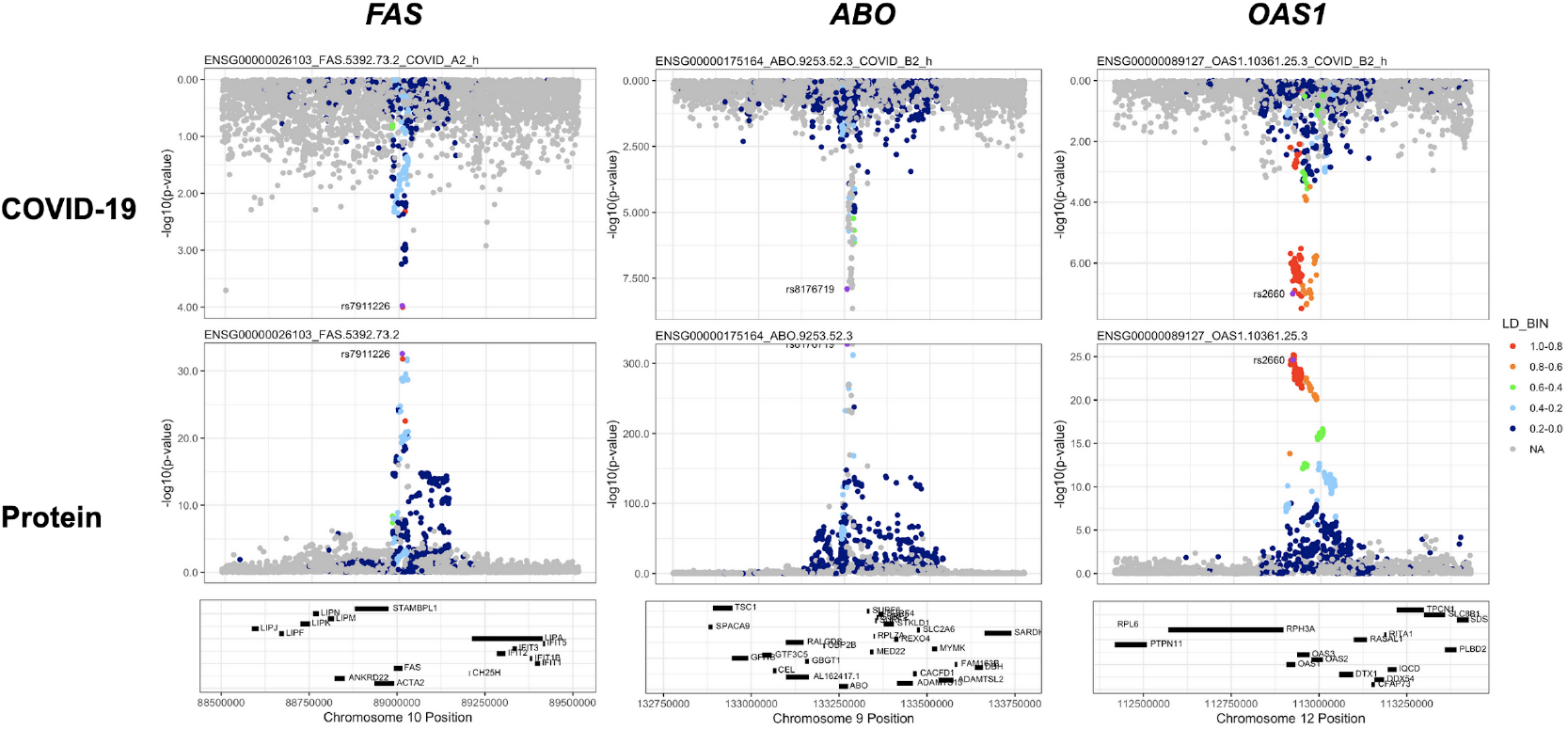
Regional association plots arranged to mirror the genetic associations of the colocalising proteins (FAS, ABO, and OAS1) with their respective COVID-19 phenotypes. The top panels represent genetic associations of the selected COVID-19 phenotypes and the bottom panels represent genetic associations of the protein from Sun et al dataset. The x-axis in each panel represents the genomic locations in or around the genes encoding FAS, ABO, and OAS1. The y-axis in each panel represents the p-value of the genetic associations.

Consistent with pan- and cis-MR findings, there was evidence of genetic colocalisation between the ABO protein and 6 out of 7 COVID-19 phenotypes, with similar MR estimates when the colocalising SNP was used to perform single-SNP cis-MR.

The coloc-first approach revealed a common genetic signal between OAS1 and COVID-19 in 2 out of 7 COVID-19 phenotypes (PP.H4 = 0.88 in COVID-19 vs. population, and 0.82 in hospitalised COVID-19 vs. population). However, the SNPs representing the common genetic signal between OAS1 and COVID-19 phenotypes (12:112919637:G:A and 12:112919388:G:A) were missense variants in *OAS1* gene or in LD with missense variants at *r*^*2*^ > 0.8, rendering their effect estimates potentially biased due to aptamer binding effects (see Methods). Despite this, the same variants have effects on gene expression (as assessed in any of several tissues curated by Open Targets Genetics^37^), which is independent of aptamer binding, suggesting that causal inference regarding OAS1 protein and COVID-19 risk may still be valid. We have, therefore, presented OAS1 estimates in **Figure 5** but flagged with an asterisk denoting the effect estimates as potentially biased.

Unlike ABO, OAS1 could not be tested using the multi-instrument MR approach due to insufficient number of valid instruments, highlighting the complementary value of the genetic colocalisation approach alongside multi-SNP MR methods. In single-SNP MR analyses, genetically predicted higher OAS1 was associated with lower risk of severe COVID-19 vs. population (OR [95% CI] per SD genetically predicted OAS1 concentrations: 0.52 [0.42, 0.65], p = 5 × 10^−9^), hospitalised COVID-19 vs. population (0.63 [0.53, 0.75], p = 1 × 10^−7^) and susceptibility to COVID-19 vs. population (0.79 [0.71, 0.87], p = 2 × 10^−6^).

Proteins that exhibited association only in one of the COVID-19 phenotypes included circulating FAS and THBS3. Genetically predicted elevated FAS (indicated by two FAS probes: FAS.9459.7.3 and FAS.5392.73.2) and THBS3 were associated with higher risk of severe COVID-19 (OR [95% CI] per SD genetically predicted FAS concentrations indicated by FAS.9459.7.3: 1.38 [1.17, 1.62], p = 1 × 10^−4^) and COVID-19 vs. lab/self-reported negative COVID-19 (OR [95% CI] per SD genetically predicted THBS3 concentrations: 1.70 [1.30, 2.22], p = 9 × 10^−5^), respectively.

### PheWAS with colocalising variants provides additional biological insights for the basis of associations of the proteins with risk of COVID-19

For the proteins with evidence of genetic colocalisation between protein and COVID-19 phenotype, we used their lead variants (or variants they tag at r^2^ > 0.6 if a lead variant was not reported in a GWAS) to identify additional associated phenotypes. At p < 1 × 10^−5^ (Bonferroni corrected for the ∼3000 phenotypes in the Open Targets Genetics portal), most of the variants exhibited associations with haematological indices, with some, like the *ABO* signal, also associated with other COVID-19 relevant phenotypes (**Table S8**). For example, the *ABO* signal was associated with monocyte count, deep vein thrombosis (DVT) and pulmonary embolism (PE). OAS1 and THBS3 variants were associated with platelet counts. For FAS, there were no additional phenotypic associations at p < 1 × 10^−5^ shown by its colocalising variant.

## Discussion

Our systematic proteome-wide MR and genetic colocalisation analysis supported several previously proposed proteins and suggested additional clinically actionable targets for COVID-19 (**Table 1**). Of particular note, we provided pan- and cis-MR evidence with strong genetic colocalisation support for the *ABO* signal for most COVID-19 phenotypes. Although the ABO protein itself is not clinically actionable, the *ABO* signal was linked to plasma concentrations of several clinically tractable targets. We demonstrated that the CD209 protein we had found to have the strongest association with this *ABO* signal has a direct interaction with the SARS-CoV-2 spike protein, providing further evidence for a plausible mechanism. Our analyses also supported the role of soluble IL-6R in hospitalised COVID-19, with evidence from pan- and cis-MR analyses but limited evidence of genetic colocalisation with hospitalised COVID-19 but supported by the recent COVID-19 clinical trials of tocilizumab (which is partially mimicked by the IL-6R instrument used in the present study). Using a proteome-wide ‘colocalisation-first’ approach, we recapitulated previously reported targets (e.g. OAS1) and uncovered additional novel proteins that may play causal roles in COVID-19 susceptibility (THBS3), or severity (FAS).

Our proteome-wide genetic colocalisation analysis prioritised soluble Fas (sFas, also known as soluble CD95) receptor protein in the very severe COVID-19 phenotype. This finding was not reported in previous proteomic MR studies of COVID-19 most likely because they only assessed evidence of shared signals for targets prioritised by an MR-first approach. Soluble Fas receptor is reported to act as a decoy receptor competing with trans-membrane Fas receptor for Fas ligand (FasL)^39^. Genetically predicted higher circulating sFas is, therefore, likely to represent effects of lower Fas-FasL signalling and, in our study, was associated with higher risk of very severe COVID-19. Fas-FasL signalling typically initiates a cascade of intracellular programs that result in cell death or apoptosis. Fas-mediated apoptosis plays a central role in T- and B-cell homeostasis^40^, preventing the emergence of autoreactive or overactive immune cells^40,41^. Excessive inflammation by hyperactive T-cells and autoantibodies were reported to underlie several cases of severe COVID-19^42^. To what extent sFas contributes to the excessive pro-inflammatory response in severe COVID-19 remains to be determined. Furthermore, Fas-mediated apoptosis of virus-infected cells is a major mechanism of resolution of viral infections^43^. Delayed apoptosis is reported to be one of the strategies exploited by SARS-CoV-2 in the early stages of infection to facilitate viral replication^43,44^. Additional insights for the role of sFas in COVID-19 can be gleaned from results of recent drug trials. For example, Fas is one of the major targets of lopinavir-ritonavir - a combination HIV protease inhibitor^45^; clinical trials of lopinavir-ritonavir failed to provide any therapeutic benefits beyond standard care in hospitalised COVID-19 patients^46^. On the other hand, clinical trials testing dexamethasone demonstrated beneficial effects on survival for COVID-19 patients who were on respiratory support^47^. In addition to its anti-inflammatory effects, dexamethasone downregulates molecules associated with decelerating apoptosis^48^, including sFas^49^. In-depth assessment of the specific role of soluble Fas in COVID-19, including whether or not it contributes to the beneficial effects of dexamethasone, is warranted in future studies.

Observational studies were the first to report differences in risk of severe COVID-19 based on ABO blood groups, although with some conflicting reports^50–52^. Genome-wide association studies of COVID-19 susceptibility have, however, consistently reported a signal in the *ABO* locus^9,13,52,53^, despite prior observations that controls used in the first published GWAS of COVID-19 (Ellinghaus et al^13^) may be over-represented for blood group O (the most common blood group) and can result in associations due to selection bias. However, in a meta-analysis of GWAS of COVID-19, the *ABO* signal remained even when Ellinghaus et al was excluded^54^. Furthermore, using these GWAS data, we had previously linked the *ABO* signal with CD209/DC-SIGN protein, clotting factors, coagulation disorders, and concentrations of IL-6^14^, all potential risk factors for COVID-19. In the present study, we build on previous work, and show consistent cis- and pan-MR associations of genetically predicted circulating ABO protein with an expanded list of COVID phenotypes which colocalise with the *ABO* signal, supporting a shared genetic signal of ABO protein and the COVID-19 phenotypes. We show that, next to the ABO protein, the *ABO* signal had the strongest association with the CD209 protein relative to other proteins and present experimental evidence of binding of CD209 with the full length spike protein of SARS-CoV-2, independently but consistent with a concurrent preprint^55^. CD209 is a receptor on monocyte-derived dendritic cells (moDCs) that was shown, before this binding interaction was known, to facilitate entry of replication-competent SARS-CoV-2 and demonstrated to switch off the type I interferon signalling pathways necessary for transcription of several antiviral genes^56^. Furthermore, CD209 gene expression was shown to be higher in immune cells (extracted from bronchoalveolar lavage fluid) in severe COVID patients than healthy controls^57^. It should be noted that the present study developed a therapeutic hypothesis for CD209 based solely on the strong evidence of association of the *ABO* signal with plasma concentrations of CD209 and evidence from the pan-MR association of CD209 with COVID-19 phenotypes (the pan-MR associations being driven mainly by trans-acting *ABO* SNPs) with no corresponding support of cis-MR or colocalisation. This suggests that while cis-MR and colocalisation analyses can support pan-MR associations of a target with disease, the lack of cis-MR or colocalisation for a target is not necessarily evidence against its therapeutic relevance.

In the present study, we also found that genetically predicted higher OAS1 - an interferon-induced broad spectrum antiviral enzyme - was associated with lower risk of both susceptibility and severity of COVID-19, consistent with findings of a recent published report^58^. A large clinical trial of systemically-administered interferons failed to show any substantial therapeutic benefits for severe COVID-19^59^. However, the strong evidence from human genetics supports reconsidering the role of interferon-based therapies in a new light, especially with respect to timing of administration (which current genetic studies are unable to provide any insights on) and route (systemic vs. nebulised)^60^.

Non-O blood group individuals generally have higher risk of deep vein thrombosis (DVT) and other coagulation disorders than O blood group individuals^61^. The *ABO* signal, which largely determines the non-O blood groups, was also associated with DVT, pulmonary embolism (PE), and higher levels of vWF and F8; vWF binds to and protects F8 from biological degradation^62^. F8 is a key protein in the intrinsic coagulation pathway that activates Factor X and induces formation of fibrin - the central component of blood clots^63^. Both DVT and PE are reported to affect almost a third of ICU-admitted COVID-19 patients^64^. While several clinical trials evaluating the efficacy of anticoagulants for severe COVID-19 are underway, the National Institute of Clinical Excellence in UK has suggested screening all hospitalised COVID-19 patients for any contraindications to anticoagulant use and offering prophylactic anticoagulation to eligible patients^65^.

We found moderate evidence for the role of IL-6 signalling in COVID-19 in agreement with a previous report^66^. However, there was ambiguous evidence of genetic colocalisation (PP.H4: 0.46). Nevertheless, there was more support for a shared genetic signal between sIL-6R and hospitalised COVID-19 than for them to be driven by independent signals (H4 / H3 = 3.6). As noted by Bovign and colleagues^66^, with some caveats, the phenotypic consistency of associations between the IL-6R genetic instrument and pharmacological effect of tocilizumab enable potential use of the IL-6R instrument to investigate therapeutic or adverse effects of tocilizumab. Although a previous report showed largely neutral effects of tocilizumab compared to placebo in hospitalised COVID-19 patients^67^, two recent trials (REMAP-CAP^68^ and RECOVERY^69^) with a longer follow-up period showed beneficial effects on survival at 90 days, consistent with the prediction of a protective effect using the tocilizumab-mimicking IL-6R genetic instrument in the present study and the previous report.

Major strengths of our study include use of both genome-wide and local genetic instruments for MR analysis, the proteome-wide genetic colocalisation tests to nominate additional proteins of therapeutic relevance, and the expanded list of COVID-19 phenotypes analysed. We showed consistency of the association of ABO with the different COVID-19 phenotypes for both instrument selection strategies. Proteome-wide colocalisation tests implicated additional proteins that likely lacked sufficient genetic instruments to be detected by the multi-instrument GSMR method. For our top-ranked association with the CD209 protein, we provide experimental evidence for a mechanism that implicates CD209 as having a potential causal role in disease pathology. Our experiments provide both direct evidence of biochemical binding between the purified spike protein of SARS-CoV-2 and CD209, and verification that this interaction occurs in live human cells. Host-directed therapies involving pathogen binding receptors have previously been developed against other infectious diseases where pathogen mutations or variants stymied more traditional approaches^70^.

Our study also has several limitations. The reliability of the MR approach depends on selection of the appropriate genetic instruments for the exposure^22^. Where proteins are the exposure, use of genetic instruments from across the genome can result in more instruments and potentially higher power to detect associations. However, inclusion of a broader set of genetic instruments for protein-MR analysis can lead to associations not mediated by the protein under investigation (i.e. horizontal pleiotropy). In these cases, use of genetic variants near or in the locus encoding the protein (cis-acting SNPs) can provide more specific estimates of risk, albeit at a potential power cost, associated with genetically predicted concentrations of the protein under investigation^22^. A key problem of the latter approach is selection of correlated genetic instruments that can lead to numerical approximation errors^71^. In the present study, we leveraged both pan- and cis-MR approaches and used an MR method (GSMR) that automates selection of near-independent genetic instruments and performs MR adjusting for any residual correlation^23^. Nevertheless, horizontal pleiotropy can also affect cis-MR analyses when different variants from the same gene region represent different biological pathways, indicated by heterogeneous effect estimates, or driven by a single variant with large effect (e.g. missense variants)^71^. To prevent selection of heterogeneous instruments and minimise selection of variants with large effects, the multi-instrument GSMR method used in the present study implements the HEIDI test which excludes genetic variants with strong or heterogeneous effects. The exclusion of missense variants with potential aptamer-binding effects is evidenced in our study, where SNPs in 96% of nominally significant protein probes associated with COVID-19 also had effects on corresponding gene expression in different tissues across gene expression datasets as curated by our portal^37^. Even while using cis-acting genetic instruments, the MR associations can be confounded due to linkage disequilibrium (LD) between cis-pQTLs and disease-associated SNPs, and this is at least partially mitigated by genetic colocalisation tests^12^. An additional issue is related to selection of COVID-19 GWAS datasets used for analyses. Most protein-MR studies have used COVID-19 phenotypes with population-based controls^10^, given their larger number of controls providing additional power to detect signals but at a cost of not being able to distinguish signals relevant to disease progression. While study designs with milder/asymptomatic cases as controls are useful to study disease progression, they are frequently under-powered and, because selection of study participants are conditioned on the outcome, are susceptible to collider-stratification bias^72^. To enable a comprehensive assessment, we used all published COVID-19 phenotypes (Oct 2020 freeze), irrespective of controls used, and, as expected, found most signals in COVID-19 phenotypes with population-based controls. For one of the targets (CD209), although we experimentally demonstrate binding of CD209 with spike protein of SARS-CoV-2, understanding the functional significance CD209 has on viral entry and any immunological relevance during infection requires further research. Finally, although we nominate several targets that may be therapeutically relevant for COVID-19, clinical trials are required for definitive assessments and to guide therapy. For example, the findings related to the *ABO* signal strongly implicated the adverse role of dysregulated coagulation in COVID-19 specifically in non-O blood group individuals; whether pre-emptive use of anticoagulants guided by blood groups can prevent severe COVID-19 are subject to findings of trials such as the on-going ACTIV-4 trial (NCT04505774)^73^.

In conclusion, we integrated genetic investigation with functional assessments of CD209, a receptor in monocyte-derived dendritic cells, and postulated that this target may convey the COVID-19 risk of the *ABO* signal. Based on proteome-wide genetic colocalisation and MR, we also prioritised sFas for more detailed investigations of its therapeutic relevance to severe COVID-19 risk.

## Data Availability

https://www.covid19hg.org/results/

## Author contributions

MAK, MG, and ID conceived the study. JH and AB harmonised all datasets. MAK performed all MR and colocalisation analysis. MC provided analytical support to carry out analyses in Google cloud virtual machines. JSh and GJW conceived and conducted the CD209 binding experiments. All authors provided valuable feedback and critical comments that informed the design and analyses in the present study.

## Supplementary Tables

Table S1: Definitions of COVID outcomes

Table S2: Summary of proteins prioritised by pan-MR

Table S3: pan-MR outcomes at p < 0.05, each association divided into cis or trans-pQTLs

Table S4: cis-MR outcomes at p < 0.05

Table S5: Evaluation of pan-MR association of protein probes that have passed the 5% FDR

Table S6: Protein-WAS (at study-specific bonferroni thresholds) of the *ABO* signal using three proteomic datasets (Sun et al, Emilsson et al, Suhre et al)

Table S7: Proteome-wide genetic colocalisation results

Table S8: PheWAS (p < 0.05) from Open Targets Genetics Portal for each colocalising variant

